# Initial clinical evidence on biperiden as antiepileptogenic after traumatic brain injury. A Randomized Clinical Trial

**DOI:** 10.1101/2023.11.10.23298341

**Authors:** Maira Licia Foresti, Eliana Garzon, Mariana Teichner de Moraes, Rafael PS Valeriano, João Paulo Santiago, Gustavo Mercenas dos Santos, Natália Mata Longo, Carla Baise, Joaquina CQF Andrade, Maria Alice Susemihl, Maria da Graça Naffah Mazzacoratti, Wellingson Silva Paiva, Almir Ferreira de Andrade, Manuel Jacobsen Teixeira, Luiz E Mello

**Author notes:** These authors contributed equally to this work (Maira Licia Foresti, Eliana Garzon). Corresponding author: Luiz Eugênio Mello, Physiology Department, Universidade Federal de São Paulo, São Paulo, Rua Pedro de Toledo 669, L3A, São Paulo-SP, Brasil, 04039-032.

## Abstract

There is currently no available drug to prevent the development of post-traumatic epilepsy. insufficient evidence regarding the effect of biperiden in preventing post-traumatic epilepsy after TBI. The combined effect of variables known to have an impact on the likelihood of developing late post-traumatic seizures and its unbalanced frequency in the different groups is an aspect to be considered and underpins the need for larger studies.

**Highlights:** Prior research suggests anticholinergics may prevent post-traumatic epilepsy

There was insufficient evidence on biperiden’s role in preventing PTE after TBI

Variable impact on post-traumatic epilepsy underscores need for broader studies

**Graphical Abstract:** 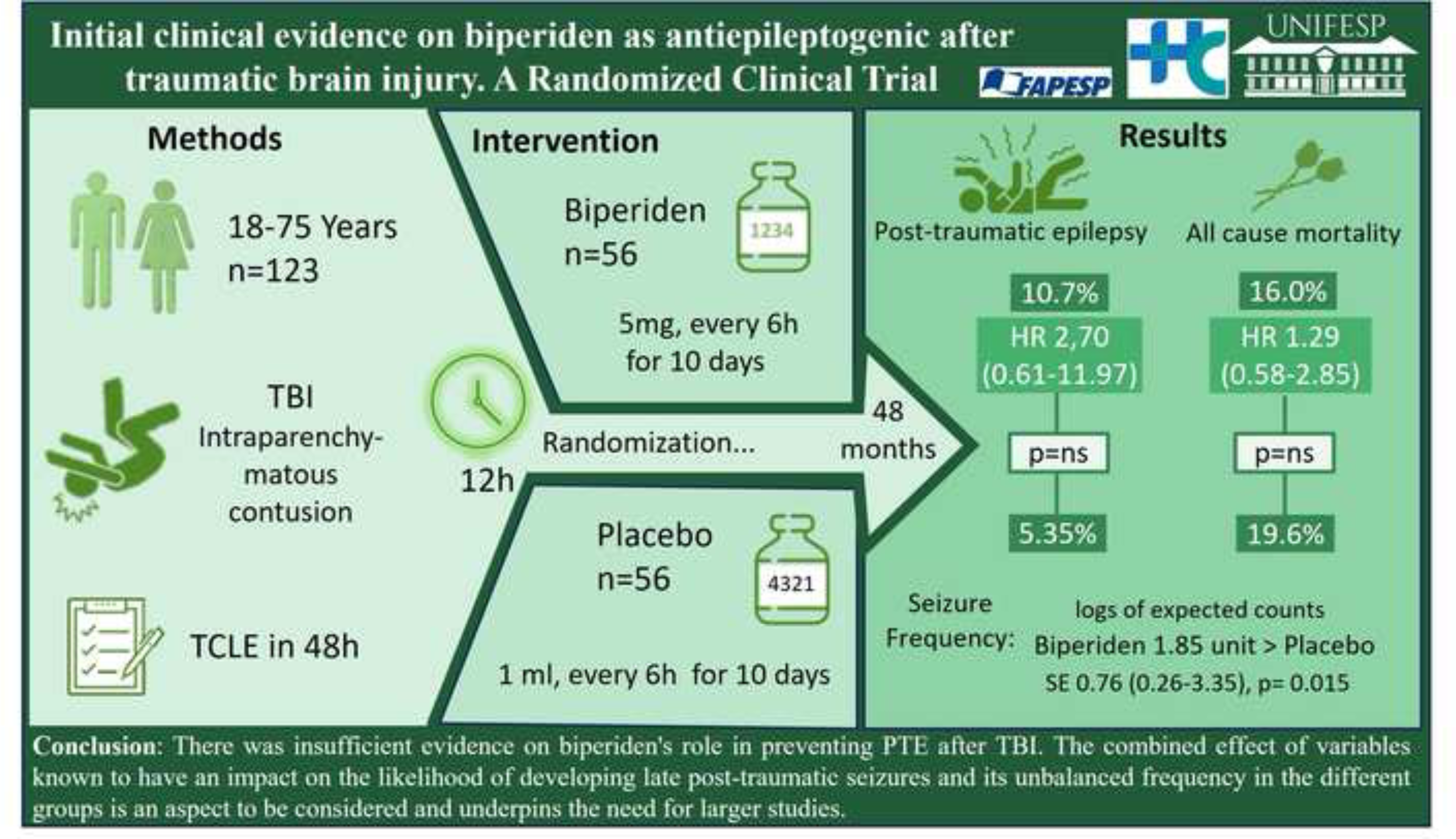

## 1. Introduction

There are no clinically available antiepileptic drugs (French et al., 2021). While this may sound weird to many, it is the consequence of recent recommendations proposed by the International League Against Epilepsy (French and Perucca, 2020). Indeed, the more appropriate terminology for the pharmacological class that was previously known as antiepileptic drugs, or AED for short, should be antiseizure medications (ASM). This change recognizes the fact that ASM are unable to suppress the underlying medical condition, that is epilepsy. In fact, ASM can be considered mainly as symptomatic drugs (French et al., 2021), which in the context of traumatic brain injury (TBI), is commonly used to treat acute symptomatic seizures (ASS, seizure occurring in the first 7 days after brain injury), but have no evidence of treatment effect on later seizures that characterize post-traumatic epilepsy (PTE) (Temkin et al., 1999, Thompson et al., 2015; Kirmani et al., 2016; DeGrauw et al., 2018; Sharma et al., 2021).

The lack of true AED in clinical use is far from being associated with a lack of research toward that end. However, considering that most PTE studies present observational research design and lack reporting the incidence of late seizures (Huo et al., 2022), together with differences in PTE definition and classification of TBI severity, among others, raises multiple barriers to the breakthrough in the field (Thompson et al., 2015). So, specific funding from many kinds of organizations, and research efforts led by groups of investigators have not been able to bring new therapeutic options (for a review see French et al., 2021).

Despite a number of potential leads provided by basic research on this topic (Sharma et al., 2021), few have progressed to clinical testing. Indeed, several hurdles must be cleared if one is to bring a finding from preclinical to clinical level. Tetrodoxin, a potent blocker of voltage-gated sodium channels has been shown to effectively influence epileptogenesis (Graber and Prince, 1999; 2004). Despite its effectiveness in vitro, it has yet to fulfill its promise as a therapeutic antiepileptogenic compound (Bucciarelli et al., 2021). Similarly, agents that are approved for use in a number of conditions in humans have been successful in affecting disease progression in animal models of epilepsy but have yet to be tested in the clinical setting as antiepileptogenic agents (Klein et al., 2020). In fact, although the safety profile of repurposed drugs is much better understood than for a new molecule, the availability of studies assessing the second use of an already existing medication, is less than it would be desired.

Here we have taken a finding of possible antiepileptogenic effects of anticholinergic drugs with no immediate application (Benassi et al., 2021) into real-world use, by associating it with biperiden, an already known and widely used drug. In addition to being a routine antiparkinsonian agent with decades of use in millions of patients worldwide (Armstrong et al., 2020), there have also been reports on the experimental use of biperiden for the treatment of depression (Kasper et al., 1981; Gillin et al., 1995). The available evidence indicates its effect on Parkinson’s to be associated with modulation of the cholinergic neurons in the striatum (Rizzi and Tan, 2017). In contrast, its potential use as an antidepressant has yet to be understood, despite some suggestions of an effect via BDNF/TrkB signaling (Zhou et al., 2017). For its purported use as an antiepileptogenic or disease modifying agent, our testing hypothesis is that of modulation of plastic phenomena (Benassi et al., 2021).

Therefore, considering that TBI leading to PTE provides the best opportunity for investigating epileptogenesis using a parallel animal/human research paradigm (Engel et al., 2019), the goal of this clinical study was to provide the initial findings of a double blind randomized, placebo-controlled study in which subjects with TBI have been treated with anticholinergic, aiming at providing a means to influence the later development of PTE.

## 2. Material and methods

To evaluate the security and efficacy of biperiden in preventing the development of epilepsy in patients that suffered TBI we performed a prospective, double blind, randomized, placebo-controlled trial. The primary outcome was change in the emergence of post-traumatic epilepsy.

### 2.1. Standard protocol approvals, registrations, and patient consents

The study protocol was approved by the Research Ethics Committee of Hospital das Clínicas da Faculdade de Medicina da Universidade de São Paulo (HC-FMUSP; CAAE n 08533513.6.2002.0068) and Universidade Federal de São Paulo (UNIFESP; CAAE n 08533513.6.1001.5505). Because of the emergency nature of TBI, we were allowed to initiate the study protocol without informed consent of patients (or relatives), which should be provided within 48h. The study was registered with ClinicalTrials.gov identifier: NCT01048138.

### 2.2. Participant recruitment and screening

Patients with acute TBI admitted at the emergency care unit (ECU) of the HC-FMUSP, between January 2018 and December 2020 were screened. Screening procedures, including standard computerized tomography (CT) scan evaluation, were performed by the resident neurosurgeon involved in the care of the patient to determine subject eligibility criteria.

Inclusion criteria were age 18-75 years; diagnosis of acute TBI admitted to the emergency unit within 12 h of the accident, regardless of the accident; brain CT scan with signs of acute intraparenchymatous contusion; signed informed consent (possibly by a relative, within 48h after inclusion). Exclusion criteria included history of epilepsy or use of ASM; previous cerebrovascular accident; malignant neoplasia and other severe comorbidities; neurodegenerative disorders; concomitant use of other anticholinergic medications; pregnancy; presence of any factor that might contraindicate the use of biperiden; participation in another clinical trial. Alcohol intoxication did not lead to exclusion of the subject.

### 2.3. General design and randomization

This trial was originally designed to enroll 132 patients, being 57 patients in each treatment arm (placebo and biperiden) and additional 18 patients to compensate eventual screening failure or follow-up loss, but recruitment and funding issues, together with the event of the SARS-CoV-2 pandemic prompted an adjustment in the study design to stop enrollment at 123 patients.

Randomization was performed using random numbers generated by the SPSS statistical package (14.0 for Windows, SPSS Inc), and was done at a 1:1 ratio of placebo and biperiden, in blocks of 6, by an unblinded investigator. The allocation was assigned by a blinded pharmacist, following the randomly generated list according to the sequential inclusion of participants admitted in the ECU and included by neurosurgeons.

### 2.4. Treatment with study medication

Once enrolled, 1 mL of biperiden (5 mg/mL; Cinetol, Cristália, Brazil) diluted in 10-50 mL of sterile saline, or placebo (1 mL of sterile 0.9% saline solution diluted in 10-50 mL of sterile saline) was intravenous administered as soon as possible within 12 h after TBI aiming at modifying the epileptogenic process. The intervention was repeated every 6 h for 10 consecutive days, until completing 40 total doses. Care providers (managing physicians and nurses) were partially blinded, given that despite lacking awareness of group assignment, minimal bottle size differences could be recognized if closely compared. Study medication was stored and dispensed by the hospital pharmacy service to the nurses engaged in the trial, which also kept the records of the distribution of medications used in this clinical trial, to provide drug accountability. Demographic and clinical data of the patients were collected, including the occurrence of acute symptomatic seizures. The incidence of already known clinical adverse events for biperiden use and other events were evaluated during the intervention period. Patients were not deprived of any medical treatment, including use of ASM (such as phenytoin), indicated for their case.

For the follow up, clinical evaluation was performed by blinded experienced epileptologists at 1, 3, 6, 12, 18 and 24 months after TBI. At each visit, any adverse events were assessed, and neurologic examination was performed, focusing on the occurrence of unprovoked epileptic seizures starting 7 days after TBI. Therefore, the PTE diagnosis was defined based on detailed clinical history that was obtained from the information of the patient and family members.

It is important to highlight that, unfortunately, due to the restrictive social measurements imposed by the SARS-CoV-2 pandemic in 2020/2021, the in person follow up assessments to verify history of seizure occurrence, and its clinical characteristics in order to classify seizures types, had to be replaced by phone calls, which continued to be performed by the same epileptologists.

The efficacy of biperiden as an antiepileptogenic drug was evaluated by comparing the development of PTE between the biperiden and placebo groups over the two-year follow-up. As part of an exploratory extended study, which results will be reported separately, acute and chronic electroencephalogram (EEG), brain magnetic resonance image (MRI) exam, genetic and behavioral data were also monitored for assessing potential mechanisms by which biperiden exerts its actions on epileptogenesis.

### 2.5. Statistical analysis

For this clinical trial, the primary outcome was incidence of post-traumatic epilepsy. In order to detect a reduction in the incidence of PTE from 23% in the placebo group to 5% in the biperiden group, with an alpha risk of 5% and power of 80%, we needed to evaluate 57 patients per group, totaling 114 patients. In anticipation of discontinuation of some patients, we planned to randomize a larger number which we estimate to be around 132 patients. As already mentioned, mainly because of the SARS-CoV-2 pandemic, this trial enrollment was terminated at 123 patients.

Baseline clinical features of the participants are presented in terms of descriptive statistics as recommended by CONSORT (Schulz et al., 2010). For continuous outcomes, means and standard deviation (SD) values are presented. For categorical data, proportions and counts are presented. For an even more precise characterization of study participants, we presented the features of age and Glasgow Coma Scale on scene (GCSoS) of the accident, as well as on hospital admission (GCSoA), both as continuous data, used at statistical analysis, and as categorized data, which is relevant for clinic interpretation.

Cox regression (Breslow method for ties) was used to assess the difference between groups on the two time-to-event outcomes (PTE and mortality). The assumption of proportional hazards was tested via Schoenfeld residuals.

Zero-inflated Poisson regression with robust standard error (Colin et al., 2009) was used to estimate the effect of the intervention on the seizure frequency (number of post-traumatic seizures), which is the secondary outcome.

We used GCSoS as an adjustment given that it is described in the literature that GCS can be a predictor of PTE (Ferguson et al., 2010; Burke et al., 2021) and, therefore, based on a clinical perspective. However, note that both unadjusted and adjusted estimates are reported to give transparency regarding the effects, especially given that our outcomes are time-to-event (Austin, 2010). Due to missing data in GCSoS and to comply with the Intention-to-treat (ITT) approach when running adjusted models, we used multiple imputations. The following measures were considered in the unrestricted models: age, sex, and group randomization. Because all those measures had non-missing values, they were used only as predictors in the unrestricted model. We imputed ten datasets and the results presented are the pooled estimates. STATA version 14 was used for all the analyses. The statistical significance level adopted was 0.05. Final analyses were performed by a blinded statistician.

The study protocol and statistical analysis plan are available in eSAP 1.

## 3. Results

A total of 122 adult patients with TBI were recruited from January 2018 to March 2020, when patient’s recruitment was suspended due to the social isolation measures imposed by the SARS-CoV-2 pandemic. A single patient was recruited in December 2021, totalizing 123 recruited patients. Of these, 59/123 patients (47.9%) were randomized for the Placebo group and 64/123 patients (52%) were randomized for the Biperiden group (Figure 1). As already pointed out, with the social restrictions measures implemented as a result of the SARS-CoV-2 pandemic, the monitoring of patients after hospital discharge was interrupted as of March 2020. After a period of reorganization, some clinical follow-ups were carried out through telephone calls, in an attempt to maintain the link between patients and the project team and to monitor the emergence of PTE. Despite being effective, there was a delay in the evaluation of many patients, especially for those completing the follow-up period in 2020/2021 pandemic. A total of 3/123 patients (2.4%) were lost in the follow-up, without completing any visits. A total of 27/123 patients (21.9%) died during the two-year follow-up (placebo n=11, biperiden n=16). In addition, 11/123 (8.9%) patients were excluded or discontinued from the study, mostly for presenting previous epilepsy, and therefore were not included in the ITT analysis. Overall, of the 82/123 (66.6%) remaining participants, a total of 61/123 (49.5%) participants (placebo n=33, biperiden n=28) completed the last study assessment, comprising the 24 months period after TCE (last appointment ranged between 23 and 45 months). Figure 1 shows the flowchart of the trial.

**Figure 1.**
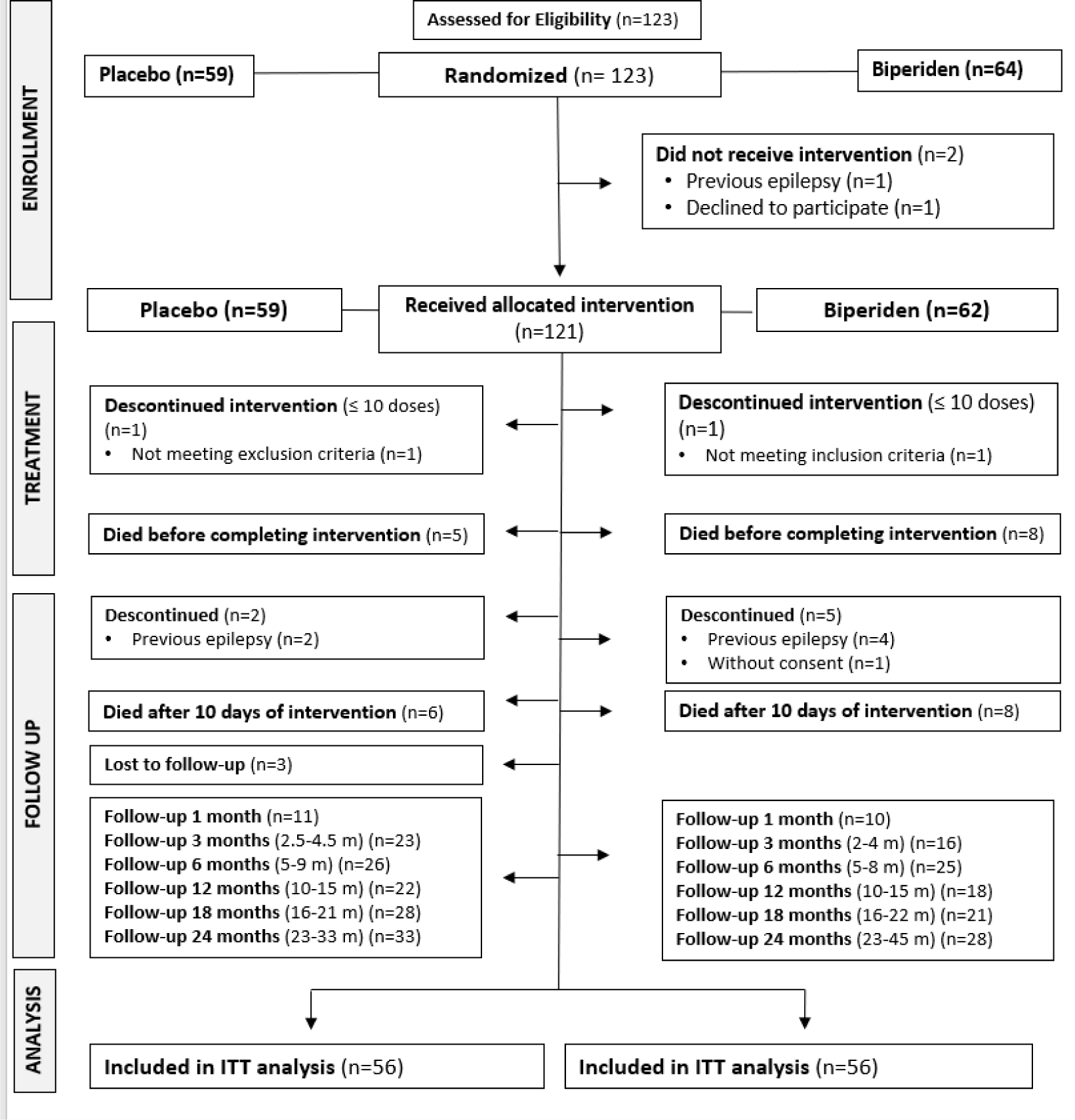
Flow diagram of randomized clinical trial for biperiden as antiepileptogenic after traumatic brain injury.

Overall, considering the 112 participants included in the ITT analysis (n=56 for each group), 19/112 (16.9%) participants were female with mean (SD) age of 49.6 (20.8) years and 93/112 (83%) participants were male, with 41.2 (16.5) years, resulting in 43.5 (17.6) mean (SD) age for the whole sample.

Table 1 and Table 2 are descriptive statistics for the participants in terms of continuous and categorical measures, respectively. It might be observed that the groups are balanced in terms of demographic and clinical features, including those related to the incidence of ASS. However, it is important to highlight that we observed some unevenness among groups when considering some clinic aspects relevant for the PTE development, e.g. given the known GCSoS (data available for 80/112 (71.4%) participants), the biperiden group had 31/41 (75.6%) participants with moderate and severe TBI, while the placebo group presented 19/39 (48.7%) participants with this TBI severity. Such distributions are more even if considering the GCSoA, but this must be considered carefully as patients could be under sedation upon arrival at the hospital. Furthermore, contributing to discrepancies among features relevant for PTE development, the number of participants with bilateral brain lesions in the biperiden group (22/56; 39.2%), was double the number of participants in the placebo group (10/56; 17.8%).

**Table 1.**
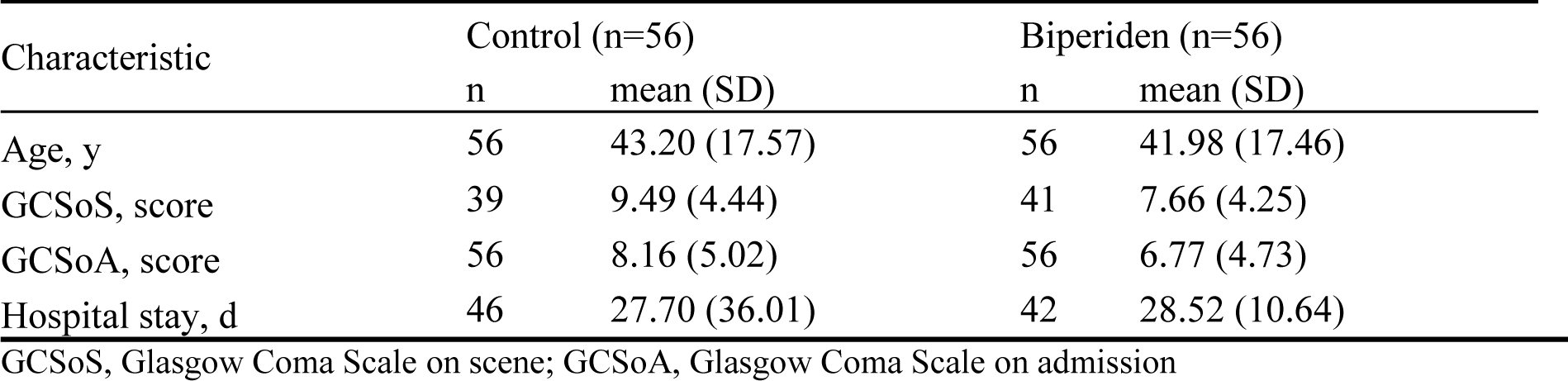
Summary statistics of the baseline characteristics of participants by allocated group Characteristic Control.

**Table 2.** Summary statistics of the baseline characteristics of participants by allocated group.

| Characteristic |  | Control (n=56) |  | Biperiden (n=56) |  |
| --- | --- | --- | --- | --- | --- |
|  |  | n | % | n | % |
| Female |  | 9 | 16.1 | 10 | 17.9 |
| Age | 16-45 | 32 | 57.1 | 35 | 62.5 |
|  | 46-88 | 24 | 42.8 | 21 | 37.5 |
| GCSoS | Mild | 20 | 35.7 | 10 | 17.8 |
|  | Moderate | 2 | 3.5 | 4 | 7.1 |
|  | Severe | 17 | 30.3 | 27 | 48.2 |
|  | Missing | 17 | 30.3 | 15 | 26.7 |
| GCSoA | Mild | 18 | 32.1 | 14 | 25.0 |
|  | Moderate | 8 | 14.2 | 3 | 5.3 |
|  | Severe | 30 | 53.3 | 39 | 69.6 |
| Acute symptomatic seizure (yes) |  | 12 | 21.4 | 10 | 17.9 |
| Neurosurgery (yes) |  | 28 | 50.0 | 35 | 62.5 |
| Phenytoin (yes) |  | 42 | 75.0 | 36 | 64.3 |
| Antibiotics (yes) |  | 43 | 76.8 | 37 | 66.1 |
| Family history of epilepsy | Yes | 27 | 48.2 | 26 | 46.4 |
|  | No | 8 | 14.3 | 4 | 7.1 |
|  | Not sure | 1 | 1.8 | 0 | 0.0 |
|  | Missing | 20 | 35.7 | 26 | 46.4 |
| *CT evaluation* |  |  |  |  |  |
| Number of lesions* | No lesion | 6 | 10.7 | 8 | 14.2 |
|  | One lesion | 27 | 48.2 | 16 | 28.5 |
|  | Two lesions | 8 | 14.2 | 13 | 23.2 |
|  | Multiple lesions | 15 | 26.7 | 17 | 30.3 |
|  | Missing** | 0 | 0 | 2 | 3.5 |
| Lateralization of lesions* | No lesion | 6 | 10.7 | 8 | 14.2 |
|  | Right hemisphere | 21 | 37.5 | 13 | 23.2 |
|  | Left hemisphere | 19 | 33.9 | 11 | 19.6 |
|  | Bilateral | 10 | 17.8 | 22 | 39.2 |
|  | Missing** | 0 | 0 | 2 | 3.5 |
| Location of lesions* | No lesion | 6 | 10.7 | 8 | 14.2 |
|  | Frontal | 30 | 53.5 | 30 | 53.5 |
|  | Temporal | 26 | 46.4 | 29 | 51.7 |
|  | Parietal | 7 | 12.5 | 6 | 10.7 |
|  | Occipital | 3 | 5.3 | 2 | 3.5 |
|  | Nucleocapsular region | 1 | 1.7 | 0 | 0.0 |
|  | Other (insula, cerebellum) | 1 | 1.7 | 1 | 1.7 |
|  | Missing** | 0 | 0 | 2 | 3.5 |
| Fracture | Yes | 31 | 55.3 | 36 | 64.2 |
|  | No | 25 | 44.6 | 18 | 32.1 |
|  | Missing** | 0 | 0 | 2 | 3.5 |
| Subarachnoid hemorrhage | Yes | 28 | 50.0 | 31 | 55.3 |
|  | No | 28 | 50.0 | 23 | 41.0 |
|  | Missing** | 0 | 0 | 2 | 3.5 |
| Subdural hematoma | Yes | 21 | 37.5 | 29 | 51.7 |
|  | No | 35 | 62.5 | 25 | 44.6 |
|  | Missing** | 0 | 0 | 2 | 3.5 |
| Epidural hematoma | Yes | 17 | 30.3 | 9 | 16.0 |
|  | No | 39 | 69.6 | 45 | 80.3 |
|  | Missing** | 0 | 0 | 2 | 3.5 |
| Midline shift | Yes | 12 | 21.4 | 13 | 23.2 |
|  | No | 44 | 78.5 | 40 | 71.4 |
|  | Missing** | 0 | 0 | 3 | 5.3 |
| Diffuse cerebral edema | Yes | 0 | 0.0 | 1 | 1.7 |
|  | No | 56 | 100.0 | 53 | 94.6 |
|  | Missing** | 0 | 0 | 2 | 3.5 |

CT, computerized tomography.
\**Lesion* refers to intraparenchymal hemorrhage and/or contusion.
\*\**Missing* refers to CT images not found for review.

Treatment compliance was assessed based on both the patient’s electronic record and the drug accountability provided by the pharmacy team. In general, patients received mean (SD) 30.95 (11.59) doses of placebo initiating 12.11 (9.99) h after TBI or mean (SD) 25.79 (14.15) doses of biperiden initiating 10.65 (5.84) h after TBI, which is far below the total of 40 planned doses. More specifically, only 31/56 (55.3%) participants received at least 30 of the total of 40 doses (75%) of biperiden. Importantly, we must consider that patients evolving to death before completing the intervention contributed to poor adherence numbers. Even so, biperiden was well tolerated for patients in the context of acute TBI. Constipation (43.9%; placebo n=28, biperiden n=26) and agitation (21.1%; placebo n=10, biperiden n=16) were the most common adverse events observed during the intervention period. Nevertheless, both events were primarily associated with prolonged bed rest and reduced levels of sedation in intensive care patients, respectively for constipation and agitation.

Death cases were mainly attributed to the TBI itself (placebo n=9, biperiden n=12); in other cases, deaths were attributed to injuries in other parts of the body (n=1 placebo), comorbidities (n=1 biperiden), sepsis (n=1 placebo, 583 days; n=1 biperiden, 82 days), malnutrition associated with major depressive disorder (n=1 biperiden, 130 days) and neurologic shock (n=1 biperiden, 214 days). One patient (placebo) had a stroke after 32 months and was not included in the mortality analysis. Cox regression models, both unadjusted and adjusted for GCSoS, showed that there is a lack of evidence regarding group differences for survival (Table 3).

For the primary outcome, 3 placebo patients and 6 patients in the biperiden group developed epilepsy during the two-year follow-up. An additional participant from the biperiden group presented alcohol withdrawal seizures 31 months after TBI, and therefore was not included on PTE analysis. Again, both Cox regression models, unadjusted and adjusted for GCSoS, showed lack of evidence regarding group differences for this outcome. Table 3 also shows the unadjusted and adjusted effects of the intervention for PTE outcome.

**Table 3.** Results of the Cox regression models.

| Outcome | Model | Coefficient | SE | P-value | HR | 95% CI |  |
| --- | --- | --- | --- | --- | --- | --- | --- |
| PTE | Unadjusted | 0.97 | 0.70 | 0.170 | 2.63 | 0.65 | 10,57 |
|  | Adjusted | 0.99 | 0.75 | 0.190 | 2.70 | 0.61 | 11,97 |
| Survival | Unadjusted | 0.45 | 0.39 | 0.248 | 1.57 | 0.73 | 3,38 |
|  | Adjusted | 0.25 | 0.40 | 0.529 | 1.29 | 0.58 | 2,85 |

PTE, post-traumatic epilepsy

For the secondary outcome, the difference in the logs of expected counts of unprovoked seizures is expected to be 2.03 unit higher for biperiden compared to placebo (robust standard error = 0.573, p-value <0.001, 95% CI = 0.912 to 3.1597). After the adjustment of GCSoS, the results are close to what was previously reported, the difference in the logs of expected counts of unprovoked seizures, is expected to be 1.857 unit higher for biperiden compared to placebo (robust standard error = 0.762, p-value 0.015, 95% CI = 0.263 to 3.351).

## 4. Discussion

This first randomized, double-blinded trial of treatment with biperiden versus placebo in patients with acute TBI shows that biperiden is safe and well-tolerated when used under the current administration regimen in critical care patients. For the primary outcome, incidence of PTE after TBI, results did not achieve significance when comparing both groups. Similar finding was achieved for survival analysis indicating lack of evidence.

For the secondary measurement, specifically late seizure frequency, this study indicated an increased count of unprovoked epileptic seizures in patients treated with biperiden during the two-year follow-up. In spite of baseline characteristics of groups being statistically similar, patients treated with biperiden tended to show more severe injuries as demonstrated by lower score at GCSoS than patients treated with placebo. Also, bilateral brain lesions were more frequent in the biperiden group. Even with discrepancies, both characteristics, severe and bilateral lesions, were already described as risk factors for PTE (Englander et al., 2003). Although statistical differences in seizure frequency were still found among groups even after the adjustment of GCSoS in the present study, we speculate that group differences in these parameters might have some clinical influence on the results.

Balancing the different experimental groups concerning trauma severity or GCSoS score would have required a distinct experimental design. Inclusion criteria in the current study required not only GCS score between 3-12 but in addition TC scan with evidence of contusion or intraparenchymal hemorrhage. Additionally, we strived for the treatment initiation time window, grounded in our laboratory evidence, to be as soon as possible. Moreover, conducting a double-blind, placebo-controlled intervention in an emergency setting entails achieving a balance that is the least disturbing for the appropriate standard treatment that patients would typically receive, alongside the requisites of the clinical investigation. In conclusion to this aspect, while we acknowledge the potential benefits of further defining patient allocation based on lesion aspects, we believe that doing so might introduce a significant burden to the study and could considerably compromise its feasibility.

Despite the variability in the incidence of PTE across different studies (Pease et al., 2022), taking into account the collective findings, we would have anticipated an occurrence rate of PTE in the current study to be around 13-37% (Armstrong et al., 1990; Santis et al., 1992; De Haltiner et al., 1997; Asikainen et al., 1999; Englander et al., 2003; Ferguson et al., 2010; Klein et al., 2012; Siig Hausted et al., 2020), but the total incidence, 8.03% (9/112), was lower than expected, especially considering the 5.35% (3/56) observed cases for the placebo group. Still, this incidence is higher than the 2.7% diagnosis of PTE described in recent studies (Liou et al., 2020; Burke et al., 2021). Considering this variability, the incidence rate should be carefully rethought to achieve validated sample size calculation through further investigation in the field.

Whereas most studies that present data on the incidence of epilepsy development after TBI have an inclusion window of 24 hours for intervention, here we restricted this interval to patients admitted in the initial 12 hours after injury. There is no reported evidence on any potential influence of the delay for medical assistance after a lesional event and the subsequent development of PTE. Yet it is conceivable that a shorter timeframe for medical intervention might increase the likelihood of a favorable outcome for most medical conditions. In pre-clinical studies, anticholinergic treatment is known to potentially modify the epileptogenic process (Bittencourt et al., 2017; Benassi et al., 2021; Sanabria et al., 2023). Specifically, biperiden suppresses spontaneous seizures in animal models of epilepsy. In addition, treatment with biperiden can delay the latency and decrease the incidence and intensity of spontaneous seizures (Bittencourt et al., 2017). Seconding our hypothesis, a recent paper suggested that scopolamine exerts antiepileptogenic/disease-modifying activity in the lithium-pilocarpine rat model, possibly involving increased remission of epilepsy as a new mechanism of disease-modification (Meller et al., 2021). Because of the robust results of biperiden and other anticholinergic drugs over suppressing the epileptogenic process in animal models, the inconclusive or even not beneficial use of biperiden in the current trial could be derived from the limitations of this study, as further discussed.

Based on experimental data from animal models we hypothesize that among the critical factors for an effectiveness for preventing PTE are the time-window for starting treatment after injury, treatment duration and drug dosage. All of those parameters have only been tested in rats and the inferred parameters employed here may need adjustments. Moreover, we faced difficulties, especially in the beginning of the study, to initiate the intervention within the scheduled 12 hours after TCE, since many patients from primary health centers arrived at our tertiary referral hospital at the limit or suppressing this restricted time-window. In addition, ensuring intervention during the first 24 h, especially while patients were unstable or undergoing surgery, as well as maintaining intervention throughout 10 consecutive days, were also challenging, given the diversity of health professionals involved in patient care (whom many times were polytraumatized), and different hospitalization units inside the hospital (which is the largest hospital complex in Latin America). Together these factors contributed to protocol deviations.

Other major issues for this study was related to patient recruitment, which can be explained considering the characteristics of a teaching hospital, with high turnover of neurosurgery medical residents and the complex nature of the trial (unstable patients in the emergency room, need to quickly check eligibility criteria of possible participant, quickly obtain a cranial CT and correctly analyze lesion images, perform the randomization and administration of the first dose within 12 h of trauma), which increased the occurrence of screening failure.

Also as a limitation, while randomization was performed using random numbers generated by statistical software, the allocation of the medication occurred by sequential inclusion of participants, which may have induced potential bias, especially considering that it was not adjusted by trauma severity. Second, we had to overcome important operational issues mainly related to the low-income characteristic of the population attended by the hospital, which added difficulty to contact and transport participants to follow-up visits, reduced operational research time, and finally the SARS-CoV-2 pandemic involving social distance policies, which limited our ability to strictly follow the designed follow-up schedule.

Indeed, the COVID-19 pandemic significantly impacted our study. Initially, we were compelled to halt patient enrollment due to the stringent measures imposed as a response to the pandemic. Various countries adopted diverse strategies to manage this severe public health crisis, and there was no clear indication of the duration for which measures like social distancing and remote work would remain in effect. Furthermore, we acknowledged the potential presence of additional variables introduced by the pandemic, which could hinder the amalgamation of data collected before and after the pandemic. Consequently, we made the determination to conclude the study. This decision not only facilitates a more thorough and comprehensive analysis of the results but also paves the way for potential adjustments in preparation for a new clinical trial.

As aforementioned, we did not find clinical evidence for our primary endpoint, the number of patients developing PTE. Unexpectedly, the use of biperiden in patients after TBI might worsen the frequency of seizures in those patients that develop PTE. Supported by these uncertain findings, we will refine planning an experimental design in a new multicenter trial to incorporate a larger sample size in an efficient time schedule; more restricted inclusion criteria limited only for patients with moderate and severe lesions as per GCS on admission; blind randomization by each center *in loco*, and for each patient (instead of sequentially); electronic case report forms using the RedCap system for data collection, reinforcement of protocol training and of structured research teams. These changes will help to constrain variability and increase study quality.

Despite decades of experimental investigation into the plastic changes that ensue after lesion events in the brain, there has been little progress using this concept into clinical testing. EpiBioS4Rx, a large collaborative effort currently being carried out is expected to yield one or more candidate antiepileptogenic treatments, as well as biomarker information, resources, expertise, and patient populations sufficient to carry out an economically feasible, full-scale clinical trial of at least one antiepileptogenic intervention (Engel, 2019). Our data directly anticipate some of the issues that should be considered when designing such trials, including those related with the time window after the precipitating injury.

## 5. Conclusion

There was insufficient evidence regarding the effect of biperiden in preventing post-traumatic epilepsy after TBI. The combined effect of variables known to have an impact on the likelihood of developing late post-traumatic seizures and its unbalanced frequency in the different groups is an aspect to be considered and underpins the need for larger studies.

## Acknowledgments

The authors thank physicians and nurses from the Neurology Department of HC/FMUSP for assistance with patient’s enrollment and data collection, especially Saul Almeida da Silva, Rodrigo Kei Kuromoto, Vitor Yamaki, Renato Vianna, Christiane Cobas, Denise Harumi Nakanishi, Claudia da Costa Leite, Dionasson Altivo Marques, Ramona Oliveira, Jenifer Fleming, Paula Vitoria Correa Ribeiro, Camilla Martins and Monica AL Moraes. We thank Rachel Riera, Rafael Leite Pacheco and Hugo Cogo Moreira for critical discussion of the study. We thank Clivandir S Silva from UNIFESP for technical assistance. We also thank Cristália (Brazil) for donating the biperiden used in this study.

## Authors’ Contribution

Maira L Foresti: Formal analysis, Funding acquisition, Methodology, Project administration, Supervision, Validation, Writing – original draft, Writing – review & editing. Eliana Garzon: Conceptualization, Formal analysis, Funding acquisition, Methodology, Supervision, Validation, Writing – original draft, Writing – review & editing. Mariana Teichner de Moraes: Investigation. Rafael PS Valeriano: Investigation. João Paulo Santiago: Investigation. Gustavo Mercenas dos Santos: Investigation. Natália Mata Longo: Investigation. Carla Baise: Investigation. Joaquina CQF Andrade: Resources, Supervision. Maria Alice Susemihl: Project administration, Supervision. Maria da Graça Naffah Mazzacoratti: Project administration, Supervision. Wellingson Silva Paiva: Resources, Supervision. Almir Ferreira de Andrade: Resources, Supervision. Manuel Jacobsen Teixeira: Resources, Supervision. Luiz E Mello: Conceptualization, Formal analysis, Funding acquisition, Methodology, Project administration, Resources, Supervision, Writing – review & editing

We confirm that we have read the Journal’s position on issues involved in ethical publication and affirm that this report is consistent with those guidelines.

## Statements

### Data availability statement

The data that support the findings of this study are available from the corresponding author upon reasonable request.

### Ethics approval statement

The study protocol was approved by the Research Ethics Committee of Hospital das Clínicas da Faculdade de Medicina da Universidade de São Paulo (HC-FMUSP; CAAE n 08533513.6.2002.0068) and Universidade Federal de São Paulo (UNIFESP; CAAE n 08533513.6.1001.5505)

### Patient consent statement

Considering the emergency nature of TBI, the informed consent of patients (or relatives) was provided withing 48h after study initiation, as approved by the Research Ethics Committee.

### Permission to reproduce material from other sources

not applicable.

### Clinical trial registration

ClinicalTrials.gov Identifier: NCT01048138

## Abbreviations

AED: Antiepileptic drugs
ASM: Antiseizure medications
TBI: Traumatic brain injury
ASS: Acute symptomatic seizures
PTE: Post-traumatic epilepsy
HC-FMUSP: Hospital das Clínicas da Faculdade de Medicina da Universidade de São Paulo
UNIFESP: Universidade Federal de São Paulo
ECU: Emergency care unit
CT: Computerized tomography
EEG: Electroencephalogram
MRI: Brain magnetic resonance image
gcsos: Glasgow Coma Scale on scene
gcsoa: Glasgow Coma Scale on hospital admission
ITT: Intention-to-treat

